# LOW BIRTH WEIGHT IS RELATED TO STUNTING INCIDENTS: INDONESIAN NUTRITION STATUS SURVEY DATA ANALYSIS

**DOI:** 10.1101/2024.06.10.24308684

**Authors:** Ine Rahmadiani, Arulita Ika Fibriana, Mahalul Azam

## Abstract

The problem of stunting in Indonesia is classified as chronic. In 2022, the prevalence of stunting in Indonesia was 21.6%, down from 24.4% in 2021. However, the percentage of stunting above 20% is still high and far from the WHO target. This study aimed to determine the relationship between birth weight and the incidence of stunting in toddlers in Indonesia. The research design was cross-sectional, using Indonesian Nutrition Status Survey Data in 2021 obtained from the Indonesian Health Ministry. The sample of this study was 20808 data. The data analysis techniques used were univariate, bivariate, and multivariate analysis. The results show that the variables that influence the incidence of stunting in toddlers in Indonesia are birth weight category (p=0.000; OR=2.720; CI=2.420-3.080), child age (p=0.000; OR=2.617; CI=2.401-2.854), gender of toddlers (p=0.000; OR=1.439; CI=1.338-1.547), mother’s education level (p=0.000; OR=1.329; CI=1.230-1.436), JKN ownership (p=0.026; OR=1.098; CI=1.011-1.191), mother’s employment status (p=0.013; OR=0.904; CI= 0.834-0.979), age of starting solids (p=0.000; OR=0.824; CI=0.767-0.886). This study found that birth weight was the most influential factor in the incidence of stunting.

## INTRODUCTION

Low birth weight (LBW) is less than 2500 grams (Cutland et al., 2017). LBW is further categorized into meager birth weight (VLBW, <1500 g) and meager birth weight (ELBW, <1000 g) (ELBW, <1000 g) (Cutland et al., 2017). Low birth weight is caused by preterm birth (PTB), intrauterine growth restriction (IUGR), or both (Cutland et al., 2017). Stunting is a physical growth disorder characterized by decreased growth velocity resulting from nutritional imbalances ((Losong & Adriani, 2017). According to the World Health Organization (WHO) Child Growth Standards, stunting is based on an index of body length over age (PB/U) or height over age (TB/U) with a z-score of less than -2 Standard Deviation.

Reducing child stunting is the first of six goals in the Global Nutrition Targets for 2025 and a key indicator in the second Sustainable Development Goal of Zero Hunger (UNSD, 2020). SDG Target 2.2 aims to end all forms of malnutrition, including achieving targets for stunting and wasting in children under 5 (United Nations, 2015). WHO has a resolution to reduce the number of stunted children by 40 percent by 2025 (United Nations, 2015). Stunting remains a public health problem in low-income countries, a condition associated with a 4.8-fold increase in mortality rates (Victora et al., 2021). By 2022, an estimated 22.3% of children under five years of age will be stunted globally (UNICEF et al., 2023). This means that 148.1 million children under 5 are too short (UNICEF et al., 2023). The gap between targets and data globally on stunting is enormous. The Joint Child Malnutrition Estimates (JME) released in 2023 show that the world is off track to achieve the World Health Assembly (WHA) target of 2025 and SDG target 2.2 (UNICEF et al., 2023).

Indonesia is one of the countries with a high stunting problem. Based on the Basic Health Research (Riskesdas) results in 2018, the prevalence of stunting among children under five was 30.8%. This figure shows a decrease in the prevalence of stunting compared to the 2013 Riskesdas results of 37.2% (Kemenkes RI, 2018). The record is the highest among regional countries in Southeast Asia (Kemenkes RI, 2018; Kusrini & Laksono, 2020). Based on data from Survey Status Gizi Indonesia (SSGI) in 2022, the prevalence of stunting in Indonesia was 21.6%. This number decreased compared to the previous year, 2021, which was 24.4%. Nevertheless, the percentage of stunting above 20% is still high and far from the WHO target. According to WHO, a public health problem can be considered chronic if stunting is more than 20 percent. This means that nationally, the problem of stunting in Indonesia is classified as chronic, especially in 14 provinces where the prevalence exceeds the national rate (*Direktorat P2PTM*, 2018). Stunting is a risk factor contributing to child mortality and a marker of gaps in human development (Vaivada et al., 2020). Stunting is a consequence of several factors often associated with poverty, including nutrition, health, sanitation, and environment (Vilcins et al., 2018). One of the most prominent risk factors for stunting is low birth weight (LBW). LBW is defined as a birth weight of less than 2500g. Apart from genetic factors, low birth weight is also an indication of premature birth or Intra Uterine Growth Retardation or IUGR.

This study aims to determine the relationship between birth weight and the incidence of stunting by considering other factors contributing to stunting among children under five (0-59 months) in Indonesia. This study used secondary data analysis SSGI 2021. Some things that distinguish this research from previous studies are the location and time of the research, which are different from previous studies, and research with the same title is still done in the scope of Indonesia.

## METHODS

### Data Source and Selection

This study used a cross-sectional design to analyze secondary data from the 2021 SSGI survey. The survey sampled 153,228 households in 14,899 census blocks from the March 2021 Susenas across 34 provinces and 514 districts/cities in Indonesia. The National Health Research and Development Agency conducted data cleaning and analysis. Stratified two-stage sampling was used to collect data. Subjects aged 0-59 months with complete information were included, totaling 20,808. Information was gathered through interviews with mothers or families, and specific data, like birth weight (LBW), were obtained from growth monitoring cards. Infants and toddlers in Indonesia are weighed monthly as part of the national growth and nutritional status monitoring program. Anthropometric measurements were performed by trained enumerators using standardized methods. The study population comprised 95,911 toddlers from the 2021 SSGI data. Simple random sampling was employed to select the 20,808 subjects used in the analysis (Figure 1).

**Figure 1.**
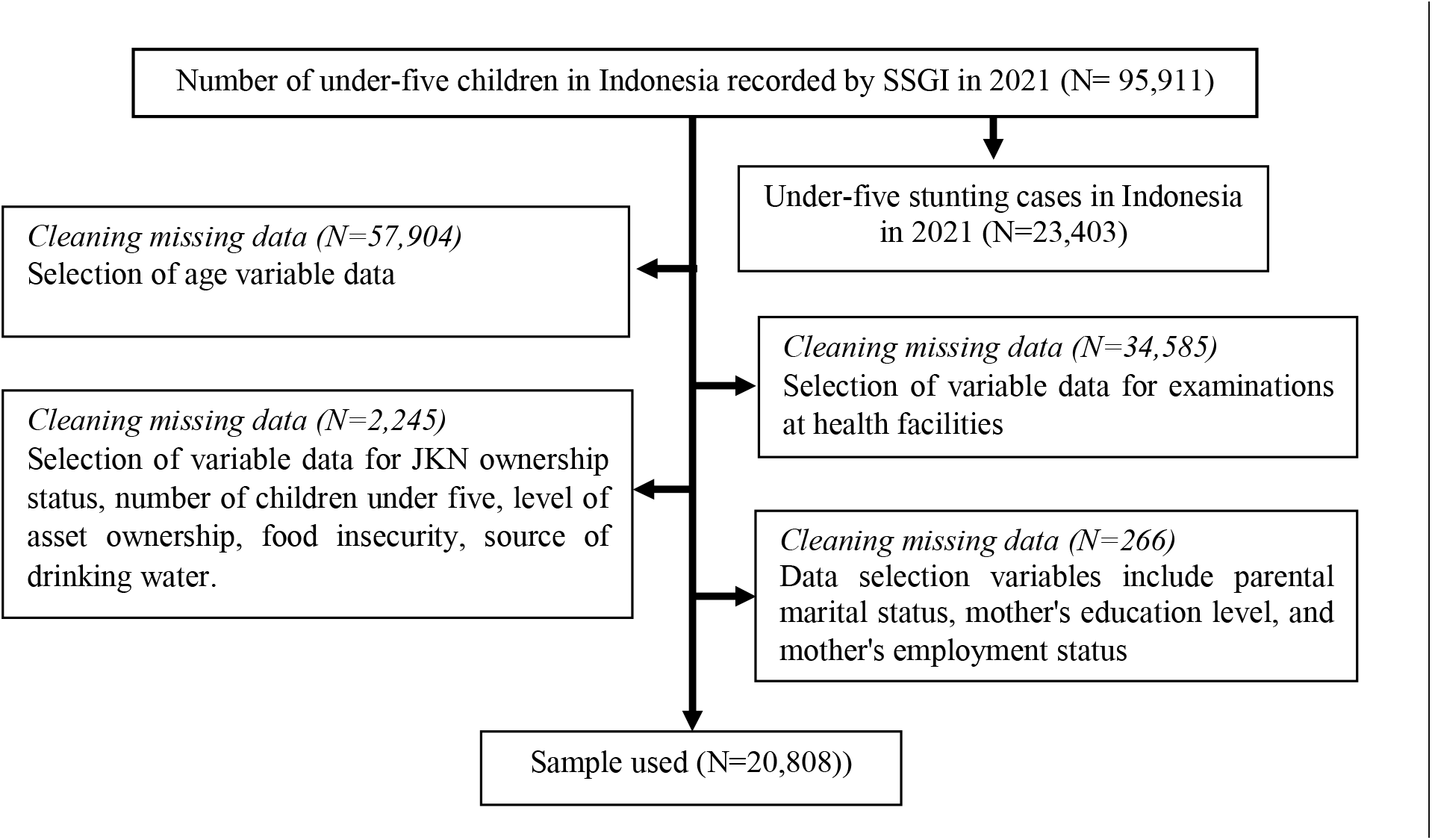
Flow of Research Sample Withdrawal

### Research Variables

The independent variables included birth weight, toddler age, age of starting solids, gender, immunization status, health facility visits, Jaminan Kesehatan Nasional (JKN) ownership, parental marital status, mother’s employment, and education level, number of toddlers, asset ownership, food insecurity, residence, and water source. The dependent variable was stunting incidence.

Birth weight was categorized as usual (≥2500 grams) or low (<2500 grams). Toddler age was calculated from birth to census year. The age of complementary feeding was in months. Gender was male or female. Immunization status was considered incomplete if not all required immunizations were received by one year of age. Vitamin A supplementation was recorded if given in the last six months before the interview. Health facility visits were noted as utilized or not. JKN ownership was categorized as yes or no. Marital status was legal or not. Mother’s employment was working or not. Education level was low (up to junior high school) or high (high school/equivalent or higher). Number of toddlers was ≤2 or >2. Asset ownership was lower/middle or upper/middle class. Food insecurity was vulnerable or not. Residence was rural or urban. The water source was protected or unprotected. Based on WHO standards, stunting was defined as a Z-score of less than -2 Standard Deviation of the median..

### Statistical Analysis

Data analysis used SPSS 25.0. Univariate analysis assessed variable distribution. Cross-tabulation and Chi-square tests (CI = 95%) examined associations between stunting and independent variables. Multiple logistic regression was used to identify significant variables (p < 0.20) with forward selection. The hypothesis was that various factors, including birth weight, toddler age, starting solids age, gender, immunization status, health facility visits, JKN ownership, parental marital status, mother’s employment and education, number of toddlers, asset ownership, food insecurity, residence, and water source influence stunting in Indonesian toddlers.

## RESULTS

The number of subjects was 20,808 children aged 0-59 months. Characteristics are described in table 1. Most subjects had a high birth weight (>2500 grams) of 93.2%. Most toddlers were> 12 months, as much as 64.5%, but the age of starting solids for toddlers was mainly at <6 months, as much as 55%. Most of the subjects were male (51.8%). Almost all subjects (100%) had incomplete primary immunization status despite utilizing existing health facilities (92%). More than half of the subjects (67.1%) did not have JKN. Most of the subjects (99.9%) had parents with married marital status, a non-working mother (68%), and a highly educated mother (50.3%). The number of children under five in the family was primarily subjects (98.6%) who had <2 children under five. Asset ownership was mostly in the middle to upper category, 51.3%. There were more toddlers with food insecurity status, 52.9%. Toddlers living in rural areas were more (54%), and almost all toddlers consumed protected drinking water (95%). Stunting was prevalent in 18.6% of subjects.

**Table 1.**
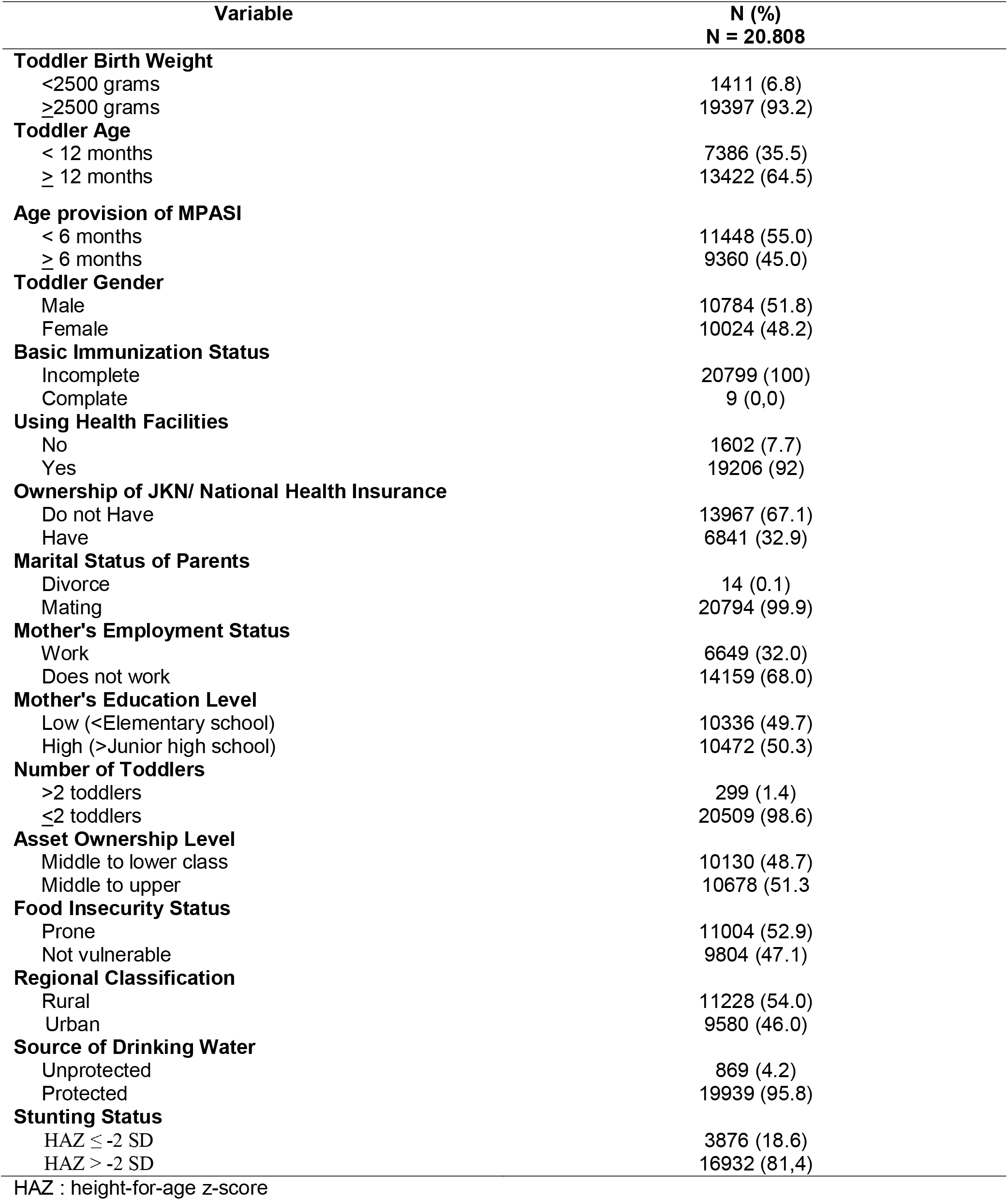
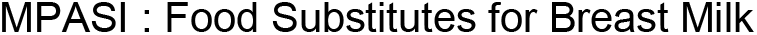
Subject Characteristics.

| Variable | N (%)<br>N = 20.808 |
| --- | --- |
| **Toddler Birth Weight** |  |
| <2500 grams | 1411 (6.8) |
| ≥2500 grams | 19397 (93.2) |
| **Toddler Age** |  |
| < 12 months | 7386 (35.5) |
| ≥ 12 months | 13422 (64.5) |
| **Age provision of MPASI** |  |
| < 6 months | 11448 (55.0) |
| ≥ 6 months | 9360 (45.0) |
| **Toddler Gender** |  |
| Male | 10784 (51.8) |
| Female | 10024 (48.2) |
| **Basic Immunization Status** |  |
| Incomplete | 20799 (100) |
| Complete | 9 (0.0) |
| **Using Health Facilities** |  |
| No | 1602 (7.7) |
| Yes | 19206 (92) |
| **Ownership of JKN/ National Health Insurance** |  |
| Do not Have | 13967 (67.1) |
| Have | 6841 (32.9) |
| **Marital Status of Parents** |  |
| Divorce | 14 (0.1) |
| Mating | 20794 (99.9) |
| **Mother's Employment Status** |  |
| Work | 6649 (32.0) |
| Does not work | 14159 (68.0) |
| **Mother's Education Level** |  |
| Low (<Elementary school) | 10336 (49.7) |
| High (>Junior high school) | 10472 (50.3) |
| **Number of Toddlers** |  |
| >2 toddlers | 299 (1.4) |
| ≤2 toddlers | 20509 (98.6) |
| **Asset Ownership Level** |  |
| Middle to lower class | 10130 (48.7) |
| Middle to upper | 10678 (51.3) |
| **Food Insecurity Status** |  |
| Prone | 11004 (52.9) |
| Not vulnerable | 9804 (47.1) |
| **Regional Classification** |  |
| Rural | 11228 (54.0) |
| Urban | 9580 (46.0) |
| **Source of Drinking Water** |  |
| Unprotected | 869 (4.2) |
| Protected | 19939 (95.8) |
| **Stunting Status** |  |
| HAZ ≤ -2 SD | 3876 (18.6) |
| HAZ > -2 SD | 16932 (81.4) |

HAZ : height-for-age z-score

Table 2 outlines the determinant of subject characteristics with stunting. LBW had the strongest significant association with stunting (OR = 2.490; CI = 2.217-2.795), followed by maternal education level (OR = 1.494; 95% CI = 1.393-1.604) and asset ownership level (OR = 1.460; 95% CI 1.361-1.566).

**Table 2.** Determinant of Stunting.

| Variable | Incidence of Toddler Stunting |  |  |  | p-value | OR | 95% CI |
| --- | --- | --- | --- | --- | --- | --- | --- |
|  | Stunting |  | Not Stunting |  |  |  |  |
|  | Σ | % | Σ | % |  |  |  |
| Toddler Birth Weight |  |  |  |  |  |  |  |
| <2500 grams | 487 | 2.3 | 924 | 4.4 | <0.001* | 2.490 | 2.217-2.795 |
| ≥2500 grams | 3389 | 16.3 | 19397 | 76.9 |  |  |  |
| Toddler Age |  |  |  |  |  |  |  |
| < 12 months | 771 | 3.7 | 6615 | 31.8 | <0.001* | 0.387 | 0.356-0.421 |
| ≥ 12 months | 3105 | 14.9 | 10317 | 49.6 |  |  |  |
| Age provision of MPASI |  |  |  |  |  |  |  |
| < 6 months | 1952 | 9.4 | 9496 | 45.6 | <0.001* | 0.794 | 0.741-0.852 |
| ≥ 6 months | 1924 | 9.2 | 7436 | 35.7 |  |  |  |
| Tooddler Gender |  |  |  |  |  |  |  |
| Male | 2255 | 10.8 | 8529 | 41.0 | <0.001* | 1.371 | 1.277-1.471 |
| Female | 1621 | 7.8 | 8403 | 40.4 |  |  |  |
| Basic Immunization Status |  |  |  |  |  |  |  |
| Incomplete | 3873 | 18.6 | 16926 | 81.3 | 0.481 | 0.458 | 0.114-1.831 |
| Complate | 3 | 0.0 | 6 | 0.0 |  |  |  |
| Using Health Facilities |  |  |  |  |  |  |  |
| No | 305 | 1.5 | 1297 | 6,2 | 0.684 | 1.030 | 0.904-1.172 |
| Yes | 3571 | 17.2 | 15635 | 75.1 |  |  |  |
| Ownership of JKN/ National Health Insurance) |  |  |  |  |  |  |  |
| Do not Have | 2707 | 13 | 11260 | 54.1 | <0.001* | 1.166 | 1.081-1.258 |
| Have | 1169 | 5.6 | 15635 | 27.3 |  |  |  |
| Marital Status of Parents |  |  |  |  |  |  |  |
| Divorce | 4 | 0.0 | 10 | 0.0 | 0.540 | 1.748 | 0.548-5.577 |
| Mating | 3872 | 18.6 | 16922 | 81.3 |  |  |  |
| Mother's Employment Status |  |  |  |  |  |  |  |
| Work | 1118 | 5.4 | 5531 | 26.6 | <0.001* | 0.836 | 0.774-0.902 |
| Does not work | 2758 | 13.3 | 11401 | 54.8 |  |  |  |
| Mother's Education Level |  |  |  |  |  |  |  |
| Low (<Elementary school) | 2240 | 10.8 | 8096 | 38.9 | <0.001* | 1.494 | 1.393-1.604 |
| High (>Junior high school) | 1636 | 7.9 | 8836 | 42.5 |  |  |  |
| Number of Toddlers |  |  |  |  |  |  |  |
| >2 toddlers | 62 | 0.3 | 237 | 1.1 | 0.385 | 1.145 | 0.864-1.518 |
| ≤2 toddlers | 3814 | 18.3 | 16695 | 80.2 |  |  |  |
| Asset Ownership Level |  |  |  |  |  |  |  |
| Middle to lower class | 2184 | 10.5 | 7946 | 38.2 | <0.001* | 1.460 | 1.361-1.566 |
| Middle to upper | 1692 | 8.1 | 8986 | 43.2 |  |  |  |
| Food Insecurity Status |  |  |  |  |  |  |  |
| Prone | 2205 | 10.6 | 8799 | 42.3 | <0.001* | 1.220 | 1.137-1.309 |
| Not vulnerable | 1671 | 8.0 | 8133 | 39.1 |  |  |  |
| **Regional Classification** |  |  |  |  |  |  |  |
| Rural | 2291 | 11.0 | 8937 | 42.9 | <0.001* | 1.293 | 1.205-1.388 |
| Urban | 1585 | 7.6 | 7995 | 38.4 |  |  |  |
| **Source of Drinking Water** |  |  |  |  |  |  |  |
| Unprotected | 168 | 0.8 | 8937 | 42.9 | 0.616 | 1.049 | 0.883-1.246 |
| Protected | 3708 | 17.8 | 7995 | 38.4 |  |  |  |

\* = *p-value* <0.05

There were ten variables included in the analysis that will be run for logistic analysis, including toddler birth weight (p = 0.000), age under five (p = 0.000), age of starting solids (p = 0.000), gender (p = 0.000), JKN ownership (p = 0.000), mother’s employment status (p = 0.000), mother’s education level (p = 0.000), asset ownership level (p = 0.000), food insecurity status (p = 0.000), regional classification (p = 0.000). Only those with significance p < 0.20 were used in the final analysis of these variables. Table 3 presents the dataset’s multivariate logistic regression analysis estimates by stunting and explanatory variables.

**Table 3.**
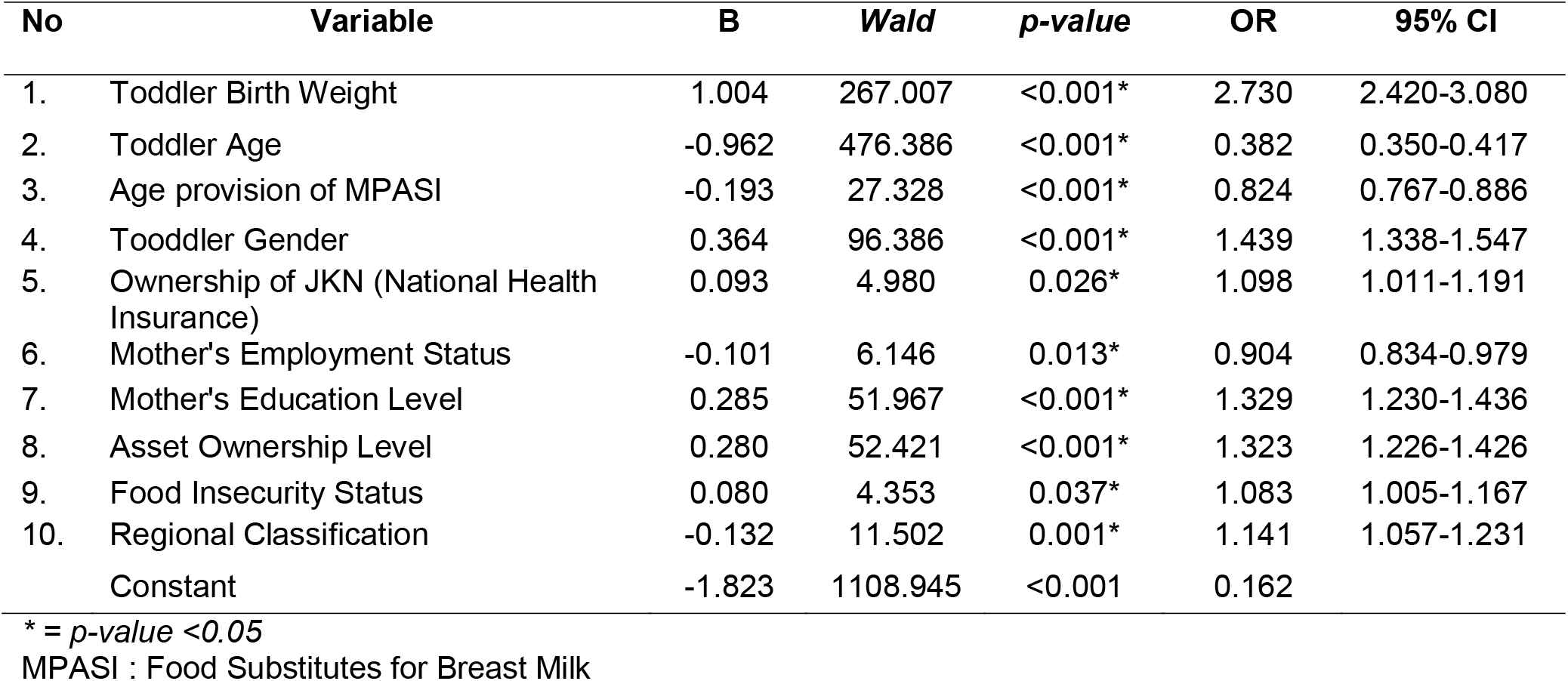
Multivariate Regression Analysis of Stunting Risk Factors.

Multivariate analysis shows that toddlers born with LBW are 2,730 times more likely to experience stunting (95% CI = 2,420-3,080) than toddlers born with average weight. Toddlers aged ≥12 months are 0.382 times more likely (95% CI = 2.401-2.854) to experience stunting than toddlers <12 months. Toddlers who start consuming MPASI ≥6 months have a 0.824 times risk of stunting compared to toddlers who consume MPASI <6 months. Toddlers who are boys are 1.439 times more likely (95% CI = 1.338-1.547) to experience stunting than toddlers who are girls. Toddlers who do not have JKN (National Health Insurance) are 1,098 times more likely to experience stunting (95% CI = 1,011-1,191) than toddlers who have JKN (National Health Insurance). Children whose mothers do not work are 0.904 times more likely (95% CI = 0.834-0.979) to experience stunting than children whose mothers work. Toddlers who have mothers with a low level of education are 1.329 times more likely to experience stunting (95% CI = 1.230-1.436) than toddlers who have mothers with a high level of education. Toddlers with families with a medium to low level of asset ownership have a 1.323 times greater chance of experiencing stunting (95% CI = 1.226-1.426) than toddlers with a medium to high level of asset ownership. Toddlers who are food insecure are 1.083 times more likely to experience stunting (95% CI = 1.005-1.167) than toddlers who are not food insecure. Toddlers who live in urban areas are 1.141 times more likely to experience stunting (95% CI = 1.057-1.231) than toddlers who live in rural areas.

## DISCUSSION

This study found that birth weight (low), gender (male), JKN ownership (not owning), maternal education level (low), asset ownership level (middle to lower), and food insecurity status (vulnerable) are risk factors for stunting in toddlers (0-59 months) in Indonesia. Meanwhile, the age of toddlers, starting solid food, the mother’s employment status, and the classification of residence are protective factors for stunting in toddlers (0-59 months) in Indonesia.

Multivariate analysis showed that low birth weight significantly affected the incidence of stunting (OR = 2,720; 95% CI = 2,420-3,080) in children under five in Indonesia. Birth weight is associated with stunting because low birth weight (BBLR) is one of the risk factors for stunting. BBLR is associated with premature birth or intrauterine growth restriction, which can affect growth achievement after birth and increase the risk of spontaneous preterm birth, increased cervical-vaginal fetal fibronectin concentrations, and lifestyle factors such as tobacco smoking (Halli et al., 2022). Studies have found that BBLR babies have less than optimal cognitive development and lag behind growth compared to normal-weight babies (Halli et al., 2022). Studies have found that BBLR babies have less than optimal cognitive development and lag behind growth compared to normal-weight babies (Halli et al., 2022). In Indonesia, BBLR is the most dominant predictor of stunting in children aged 12-23 months (Aryastami et al., 2017). In India, the study found that low birth-weight babies have a 19% higher chance of stunting than normal birth-weight babies (Halli et al., 2022). Low birth-weight babies are more at risk of stunting than normal-weight babies because they are at higher risk of not receiving enough nutrition during critical developmental periods. This can lead to impaired cognitive development and increased susceptibility to chronic diseases later in life (Halli et al., 2022). In addition, BBLR babies are more likely to be born prematurely, which further increases the risk of stunting (Halli et al., 2022). BBLR is directly related to pediatric morbidity, related to inadequate weight and height growth, neuropsychomotor developmental disorders, and an increased risk of chronic non-communicable diseases such as obesity, systemic arterial hypertension, type 2 diabetes mellitus, and cardiovascular disease in the future (Kuhn-Santos et al., 2019). Children with low birth weight (under 2,500 grams) are at higher risk of malnutrition, infections, and degenerative diseases. Malnutrition and infections can negatively impact growth and development and increase a child’s morbidity later in life (Manggala et al., 2018).

The 2018 Global Nutrition Report showed that 22.2% of children under the age of five worldwide are stunted, with Africa having the highest prevalence at 58.7% (Afework et al., 2021). Similarly, a study in Timor-Leste found that almost 40% of children under five are stunted (Nomura et al., 2023). Toddlers older than 12 months are related to stunting compared to toddlers less than 12 months old due to several factors. Toddlers over 12 months are more likely to experience nutrient deficiencies, especially essential nutrients such as iron, zinc, and calcium, essential for growth and development. This deficiency can lead to stunting (Nugraheni et al., 2023). Around 12 months, children usually begin to wean out of breast milk and switch to complementary foods. If the food is of poor quality or inadequate, this can lead to stunting (Elsa Octa Aditia et al., 2023). Toddlers over 12 months are more susceptible to infections, which can further exacerbate stunting. Infections can lead to nutrient malabsorption, further interfering with growth and development (Elsa Octa Aditia et al., 2023; Nugraheni et al., 2023). Maternal factors such as maternal height, duration of breastfeeding, and socioeconomic status also play an essential role in the occurrence of stunting. Toddlers over 12 months are more likely to be stunted if their mother has a lower height or less than optimal breastfeeding practices (Titaley et al., 2013). Environmental factors such as access to clean water, sanitation, and hygiene can also impact stunting. Toddlers over 12 months may be more susceptible to environmental factors that contribute to stunting, such as poor living conditions or lack of access to health services (Mufida Wulan Sari et al., 2023; Tyas & Setyonaluri, 2022). Combining these factors increases the likelihood of stunting in toddlers over 12 months compared to toddlers under 12 months.

The delay in starting complementary foods (for breast milk) is related to stunting because it can result in a lack of nutrient intake for children’s growth and development. Several studies support this. Children who receive complementary foods at an older age have a higher risk of stunting. Research in Sedayu Regency found that children who started MP-ASI after six months of age had a 2.8 times higher risk of stunting than children who started it early (Nilandita, 2018). Late supplementation can result in inadequate nutrient intake, essential for growth and development. After six months, breast milk alone may not provide enough energy and nutrients for the child’s needs (Soliman et al., 2021). Exclusive breastfeeding for too long can also delay the provision of complementary foods, thereby causing insufficient nutritional intake and increasing the risk of stunting. After six months, breast milk alone may not provide enough energy and nutrients for the child’s needs (Soliman et al., 2021).

Gender is one of the predictors of stunting because there are biological and socioeconomic factors that make boys at higher risk of stunting than girls. Biologically, boys are more susceptible to infectious diseases and exhibit higher biological fragility in the first year of life, which can increase the risk of stunting (Samuel et al., 2022). Socio-economically, boys may have lower access to adequate nutrition due to gender-based inequalities, such as girls being prioritized for food in some households (Thurstans et al., 2020). This can result in boys being more likely than girls to be malnourished and stunted (Thurstans et al., 2020)

National Health Insurance (JKN) coverage is associated with stunted toddlers because JKN increases the likelihood of accessing health services, improves overall health, and reduces the risk of stunting. Without JKN, children may not receive the necessary health services, increasing the risk of stunting due to a lack of adequate nutrition and health services. A study in West Sulawesi, Indonesia, found that only 36.2% of stunted children under five have JKN, while the use of JKN for stunted children is 26% (Riestiyowati & Rustam, 2022). This shows that only a few with JKN use it effectively to overcome stunting. In addition, a study on the impact of community-based health insurance on stunting in children found that one year of household participation in community-based health insurance was associated with a 4.3 percentage point reduction in the likelihood of stunting stunting (Aziz et al., 2022; Chen & Chu, 2019). This further supports the idea that health insurance can play an essential role in reducing stunting in children. In addition, research shows that maternal health insurance subscriptions affect stunting and underweight in children through maternal health (Koomson, 2022).This shows that maternal health, which can be improved through access to health services, can positively impact children’s health and reduce the risk of stunting.

Mothers who do not work have a greater risk of stunting than working mothers due to several factors. Non-working mothers may need more access to resources, increasing the likelihood of their child being malnourished. This can result in stunting because children need enough nutrition for their growth and development (Adla et al., 2022; Supadmi et al., 2024; Win et al., 2022). Non-working mothers may experience reduced access to health services, including vaccinations and preventive care, which can contribute to stunting (Supadmi et al., 2024; Menang et al., 2022). Households with non-working mothers may have more limited access to clean water and proper sanitation, increasing the risk of infection and malnutrition, both of which can lead to stunting (Independent DKK, 2024).

The low level of maternal education can be associated with the incidence of stunting in children under five due to several factors. Maternal education is associated with better knowledge and understanding of child nutrition and health. Women with higher levels of education tend to be more aware of the importance of a balanced diet, the frequency of meals, and the amount of food given to their children, which can contribute to better child nutrition. On the other hand, mothers with low levels of education may have less knowledge about child nutrition, leading to inadequate feeding practices and poor child growth (Ahmed et al., 2022; Javid & Pu, 2020; Rahayuwati dkk., 2023; Vollmer et al., 2017). Women with higher levels of education have a more significant opportunity for better employment opportunities, which in turn can improve the family’s economic condition. This can result in better access to food, health services, and other essential resources for a child’s growth and development.

Conversely, mothers with lower levels of education may have limited employment opportunities, resulting in lower incomes and lower access to resources for their children’s nutrition and health (Rahayuwati et al., 2023). Women with higher levels of education tend to have better parenting skills, which include understanding child development, providing a stimulating environment, and responding appropriately to their child’s needs. These factors can contribute to better child growth and development, including reducing the risk of stunting (Vollmer et al., 2017).

Medium to low asset ownership levels are associated with stunting toddlers due to the influence of socioeconomic factors on children’s nutrition and health. Studies have shown that an increase in asset index scores, which measure the socioeconomic status of households, is associated with improved linear growth outcomes and decreased stunting in children (Vaivada et al., 2020). In addition, women’s autonomy in asset ownership and decision-making reduces anemia and the incidence of anemia and stunting in children (Christian et al., 2023). This suggests that higher levels of asset ownership can result in better nutritional outcomes for children under five (Chirande et al., 2015).

Food insecurity is associated with stunting in children under five because food insecurity can cause malnutrition, which is a significant risk factor for stunting in children. Studies have shown that food insecurity is significantly linked to stunting in children. For example, a study published in the Journal of Nutrition found that food insecurity strongly predicts stunting (Mahmudiono et al., 2018). Another study published in the Journal of Public Health found that severe food insecurity was significantly linked to stunting in children (Gassara & Chen, 2021). Other research also states that very food-insecure households are associated with stunting (Afework et al., 2021).

Urban areas can contribute to the incidence of stunting in toddlers (0-59 months). Other research supports this finding. Urban environments, particularly in Indonesia, can contribute to the risk of stunting in toddlers through various factors, including socioeconomic and parental characteristics (Hamida Sari Batubara et al., 2023; Siswati et al., 2020). Housing is associated with stunting in children under five because inadequate housing conditions can lead to poor nutrition, water, and sanitation problems, and exposure to harmful environmental factors (Tasnim et al., 2017; Tustingi et al., 2020; Yani et al., 2023).

The weaknesses of this study include not considering other variables that contribute to the incidence of stunting and have a large missing value in several variables.

## CONCLUSION

This study found that birth weight (low), gender (male), JKN ownership (not owning), maternal education level (low), asset ownership level (middle to lower), and food insecurity status (vulnerable) are risk factors for stunting in toddlers (0-59 months) in Indonesia. Meanwhile, the age of toddlers, the age of starting solid food, the mother’s employment status, and the classification of residence are protective factors for stunting in toddlers (0-59 months) in Indonesia. The variable that most affects the incidence of stunting in children under five in Indonesia is birth weight, where toddlers with low birth weight (<2500 grams) are 2,730 times more likely to be stunted than toddlers with an average birth weight (>2500 grams). The weakness of this study is that it does not consider other variables that contribute to the incidence of stunting. Therefore, suggestions for future research are expected to include the variables considered and other potential factors that may affect the incidence of stunting, such as environmental factors and other parenting factors. In addition, more research is needed to evaluate effective interventions to prevent and reduce stunting, including integrated nutrition programs and public health interventions.

## Data Availability

All data produced in the present study are available upon reasonable request to the authors

